# Assessment of service availability and Infection prevention measures in hospitals of Nepal during the transition phase of COVID-19 case surge

**DOI:** 10.1101/2020.05.13.20097675

**Authors:** Suraj Bhattarai, Jaya Dhungana, Tim Ensor, Uttam Babu Shrestha

**Author notes:** **Corresponding author:** Dr. Suraj Bhattarai; Global health programme, Global Institute for Interdisciplinary Studies (GIIS), Kathmandu, Nepal;,; GPO Box 24560. www.thegiis.org.

## Abstract

As with other coronavirus-affected countries, Nepal’s medical fraternity also expressed concerns regarding the government’s public health strategies and hospital readiness in response to upgoing case surge. To gauge such response, we assessed service availability and Infection prevention and control (IPC) status in 110 hospitals situated across seven provinces. An electronic survey was sent out to the frontline clinicians working on those hospitals between 24^th^ March and 7^th^ April 2020; one response per hospital was analyzed. Hospitals were divided into small, medium, and large based on the total number of beds (small:≤15; medium:16-50; large:>50), and further categorized into public, private, and mixed based on the ownership. Out of 110 hospitals, 81% (22/27) of small, 39% (11/28) of medium, and 33% (18/55) of large hospitals had not allocated isolation beds for COVID-19 suspects. All small, majority of medium (89%; 25/28), and 50% of large hospitals did not have a functional intensive care unit (ICU) at the time of study. Nasopharyngeal (NP)/throat swab kits were available in one-third (35/110), whereas viral transport media (VTM), portable fridge box, and refrigerator were available in one-fifth (20%) of hospitals. Only one hospital (large/tertiary) had a functional PCR machine. Except for General practitioners, other health cadres—crucial during pandemics, were low in number. On IPC measures, the supplies of simple face masks, gloves and hand sanitizers were adequate in the majority of hospitals, however, N95-respirators, Filter masks, and PPE-suits were grossly lacking. Government’s COVID-19 support was unevenly distributed across provinces; health facilities in Province 2, Gandaki, and Province 5 received fewer resources than others. Our findings alert the Nepalese and other governments to act early and proactively during health emergencies and not wait until the disease disrupts their health systems. Other countries with similar economy levels may undertake similar surveys to measure and improve their pandemic response.

## 1. BACKGROUND

The World Health Organization (WHO) declared COVID-19 as Public Health Emergency of International Concern (PHEIC) on 30^th^ January 2020; on 11^th^ March, it was declared a global pandemic.(1) As of 12^th^ May 2020, the disease has spread to 215 countries, areas and territories, infecting four million people and causing over 279,000 deaths.(2) The increasing trend of new cases and deaths is further worsening COVID-19 situation globally. To minimize the burden, the global scientists have urged for pre-emptive measures to prevent COVID-19 outbreaks among the most vulnerable population of the world living in the vulnerable regions.(3)(4)

In Nepal, the first case of COVID-19 disease was confirmed on 24^th^ January 2020.(5) The second case was detected on 23^rd^ March, after a two-month gap. It was that day the government decided to impose a nationwide lockdown as a measure to contain virus spread.(6) However, by the end of March, Nepal Public Health Laboratory (NPHL) located within the premises of Department of Health Sciences (DoHS), Teku was the only authorized lab for polymerase chain reaction (PCR) tests. Even with the gradual expansion of PCR-based testing outside Kathmandu, the government had tested less than 1,000 specimens, with detection of nine cases, by 7^th^ of April. The number of cases reached 217 on 12^th^ May 2020.(6)

The medical fraternity of Nepal expressed their concerns regarding the government’s public health strategies and hospital readiness in response to the potential outbreak of COVID-19 in the country. The clinicians and public health experts were particularly concerned about the government’s weak preparation and unclear strategies around molecular testing, contact tracing, medical procurement, resource allocation, infection prevention measures, human resources and training, risk communication, and management of suspected or confirmed cases. (7) The nation’s case finding strategy was heavily criticized by the experts calling it ‘sheer undertesting’ for the country with 29 million people.(8)

The Government of Nepal designated 25 large (tertiary care) hospitals across all provinces as COVID-19 treatment centers, but there were no stringent guidelines for measuring pandemic readiness and response across the range of facilities.(6) With the decision of the High Level Coordination Committee (HLCC) for the Prevention and Control of COVID-19 (17^th^ March) to add 115 ICU and 1,000 isolation beds in the health facilities of Kathmandu, and set up a total of 120 ICU beds in other provinces, the overall status of pandemic preparedness and the capacity of national health systems were unknown.(9)

The WHO released ‘Hospital Readiness Checklist for COVID-19’—a useful tool for the assessment of pandemic response practices in the health facilities worldwide.(10) Similarly, the United Nations Nepal recently published ‘Preparedness and Response Plan for Nepal (NPRP)’ that highlighted key interventions that need to be implemented by the key players taking into account the projected caseload of 1,500 infected people and 150,000 collaterally affected population.(11)

The present study aimed to understand the ground status of COVID-19 services and infection prevention and how it varied across small, medium, and large hospitals. Besides few case reports and opinion pieces found in the literature, this is perhaps the first study that collected COVID-19-related primary data in Nepal from the clinical-public health point of view.

## 2. METHODS

The clinicians working at the frontline managing either suspected or confirmed COVID-19 patients in various hospitals of Nepal were included in this study—they were medical interns, medical officers, postgraduate residents, and specialists registered with Nepal Medical Council. The study participants worked in small, medium or large hospital, categorized for this study on the basis of bed capacity; small hospital: 15 or less beds (primary health centers or PHCs), medium hospital: 15-50 beds (district or community hospitals, polyclinics), and large hospital: >50 beds (zonal or provincial hospitals, tertiary centers, medical teaching hospitals).

The list of small, medium, and large hospitals in each of the seven provinces was prepared after reviewing the national health data. According to the Annual Health Report 2017/2018, there are total 2,145 health facilities in Nepal: 191 in Province 1; 214 in Province 2; 1,239 in Bagmati; 139 in Gandaki; 218 in Province 5; 71 in Karnali; 73 in Sudurpaschim. Among them, 125 are public facilities, 198 are primary health centers, and 1,822 are non-public facilities. (12)

Non-probability purposive sampling method was used to enlist potential respondents (clinicians) working in various hospitals. Sample size was calculated by applying Cochrane’s formula for a finite population (hospitals in the present study) i.e. *n=N/[1+N(e)^2^]*, where, N is total number of health facilities in Nepal, and *e* is permissible error taken as 10%. Uneven distribution of health facilities across the provinces, especially abundant in Bagmati and the scarce in others, were accounted for. Hence, a total of 110 hospitals were selected for the study: 19 in Province 1; ten in Province 2; 33 in Bagmati; 21 in Gandaki; 14 in Province 5; eight in Karnali; five in Sudurpaschim. We retained one response per hospital. For more than one responses received from the same hospital, the response of the senior-most clinician was used for analysis. Seniority was determined by the respondent’s qualification, experience, and leadership role in the hospital.

A semi-structured questionnaire was prepared after reviewing hospital readiness and IPC guidelines recommended by the WHO and Nepal government.(10)(11) For data collection, a standard survey link developed using Google forms was sent out to potential respondents by electronic (online) methods. All responses were checked for completeness and quality. We generated frequency tables out of coded data using SPSS 16.0 and prepared maps using ArcGIS.

Ethical approval to conduct this study was obtained from Nepal Health Research Council (*Ref: NHRC-ERB Reg. No. 267/2020P*). Informed consent was obtained from each study participant on the digital/electronic format. They were asked to provide information about their working district and hospital type (primary, district/community, tertiary); however, to maintain the privacy and confidentiality throughout the study, they were not obliged to provide the hospital name and other personal information. All personal identifiers were removed during data analysis and report writing.

## 3. RESULTS

### 3.1. Respondents and type of hospitals

This study covered 52 out of 77 districts of all seven provinces of Nepal (Figure 1). We received highest response from Kathmandu (15) followed by Kaski (11) districts and only one response from 31 districts. Out of 110 respondents from 110 hospitals, 65 (59%) were specialists, 34% were postgraduate trainees, and 7% were medical officers or interns. The respondents worked in small (27), medium (28), and large (55) hospitals across the country, and represented public (65), private (31), and public-private mixed (14) facilities (Figure 2).

**Figure 1.**
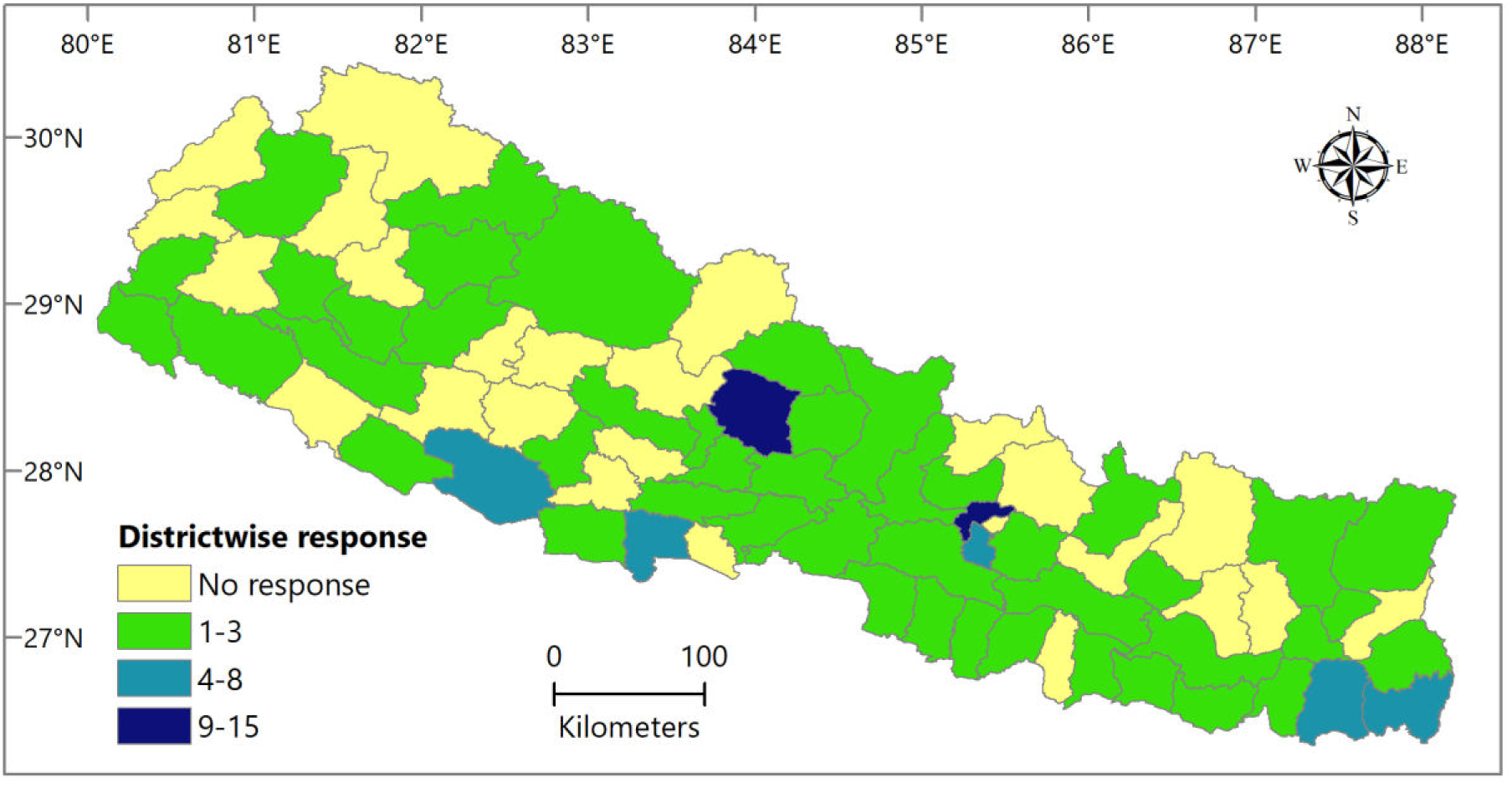
Number of responses (hospitals) under study by district.

**Figure 2.**
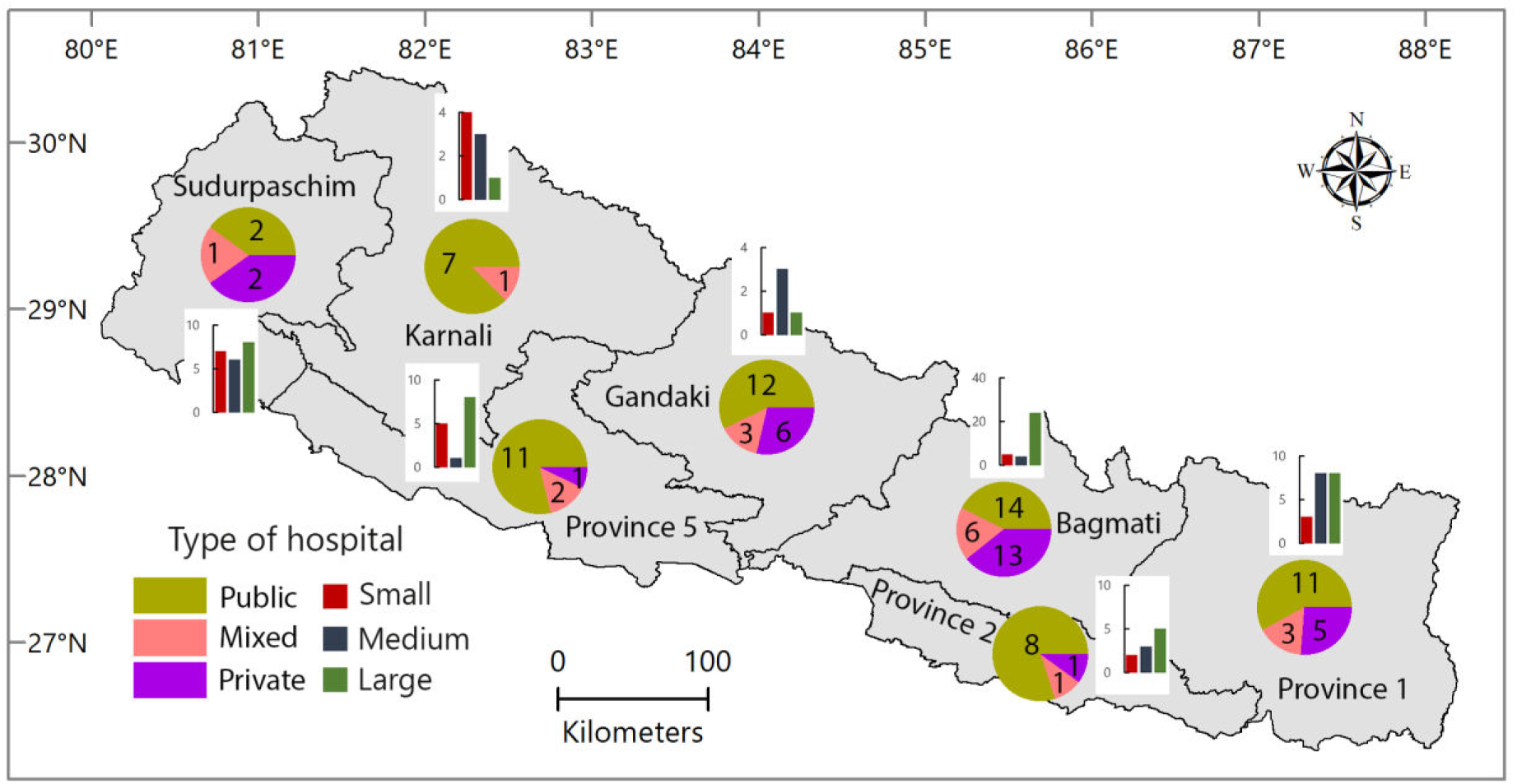
Number of hospitals under study, by province and hospital type (N=110).

### 3.2. Case surge capacity

The patient flow in pre-lockdown period was <50 per day in the majority of small hospitals (17/27), 51-200 per day in medium (20/28), and >200 per day in large hospitals (37/55) (Figure 3A). A significant number of patients with respiratory symptoms visited the facilities (up to 50 per day). However, fifteen large hospitals reported >50 patients with COIVID- related symptoms each day, with three hospitals reporting >200 daily (Figure 3B).

**Figure 3.**
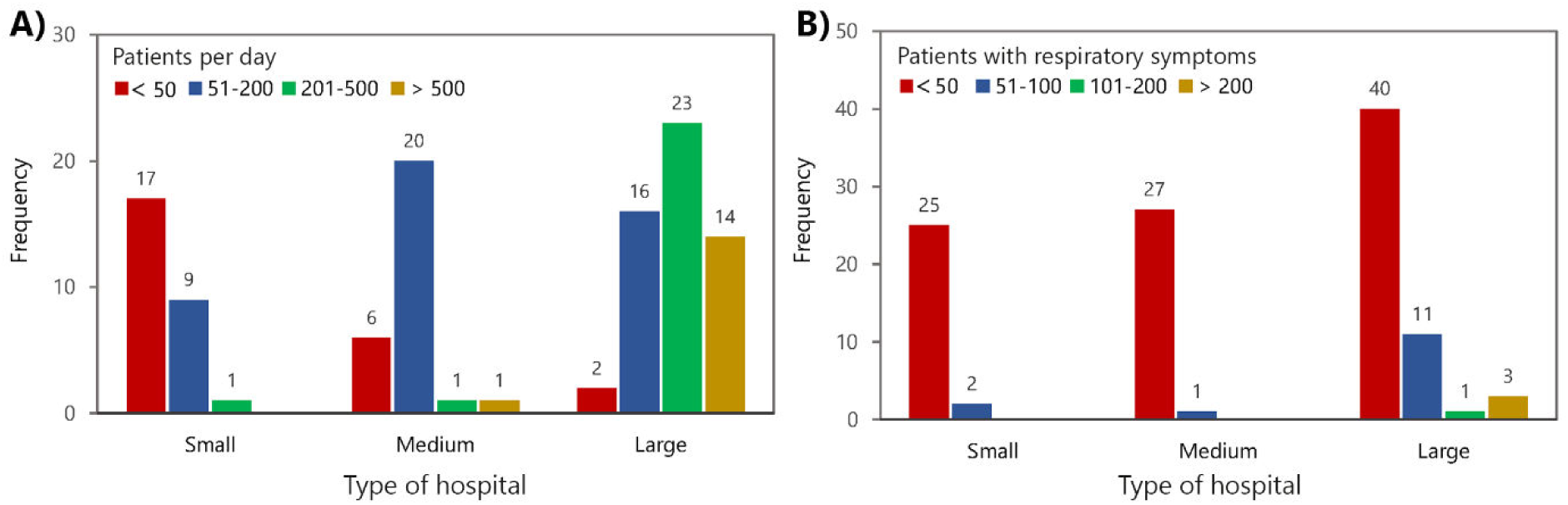
Average patient flow (A) and patients with respiratory (COVID-like) symptoms (B) attending small, medium, and large hospitals per day (N=110).

### 3.3. Special health services for COVID-19

Majority of the small hospitals (81%; 22/27) had no isolation beds for suspected COVID-19 patients. Of 28 medium hospitals, 17 had allocated few isolation beds (less than 10). Among 55 large hospitals, 21 (38%) had <10 beds, ten (18%) had 10-20 beds, and six (11%) had >20 beds designated for the isolation of COVID-19 suspected cases. One-third of large hospitals had not allocated a single bed for such patients (Figure 4A).

None of the small hospitals had an intensive care unit (ICU). Majority of the medium (25/28), and almost half of the large hospitals did not have a functional ICU bed at the time of the study. Out of 24 large hospitals that had functional ICU beds, 17 (71%) had less than 5, ten had 6-15, four had 16-25, and three had more than 25 beds that could be used for COVID-19 patients whenever there was a need (Figure 4B).

**Figure 4.**
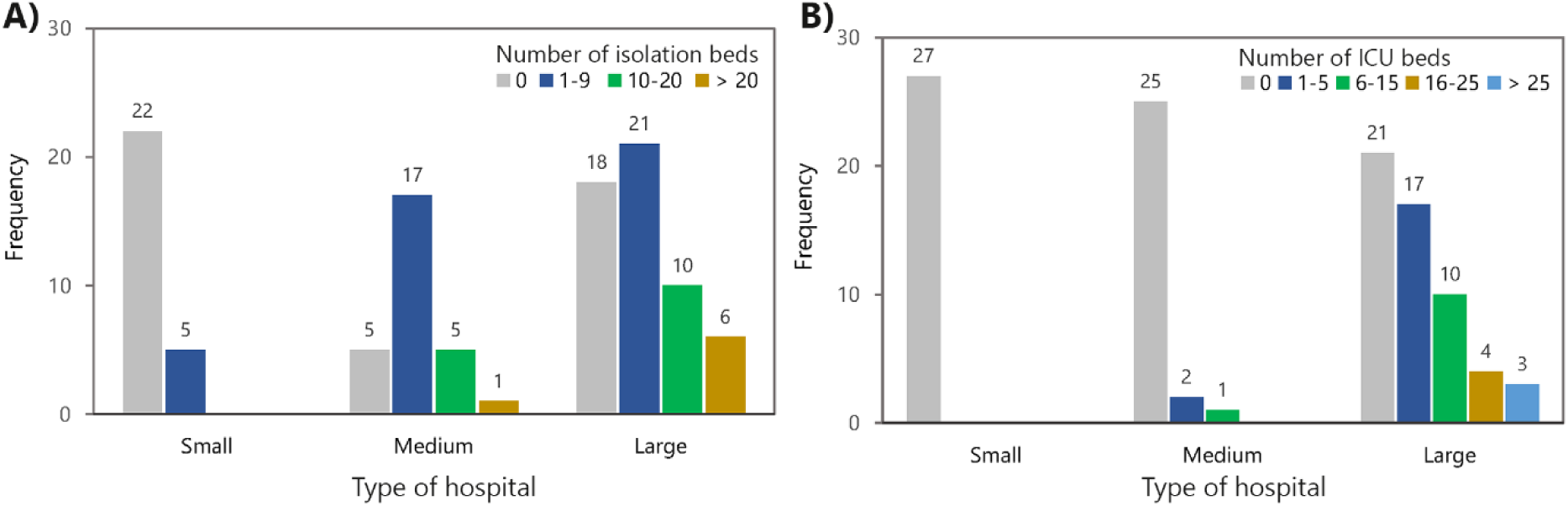
Number of isolation beds (A) and ICU beds (B) available for COVID-suspected cases in small, medium, and large hospitals (N=110).

### 3.4. Laboratory services

Majority of the small (85%; 23/27) and more than half of the medium (54%; 15/28) hospitals were found to have no capacity to collect patient’s respiratory specimens. Four medium hospitals had partially adequate and four had adequate capacity to collect samples (all public facilities). Among 39 (71%) large hospitals that started collecting samples, 24 had inadequate, eight had partially adequate, and only seven had adequate capacity (Supplementary table 1).

**Supplementary Table 1.** Availability of laboratory services, equipment, and personnel in small, medium, and large hospitals

|  |  | Type of Hospital (N=110) |  |  |  |  |  |  |  |  |
| --- | --- | --- | --- | --- | --- | --- | --- | --- | --- | --- |
|  |  | Small (n=27) |  |  | Medium (n=28) |  |  | Large (n=55) |  |  |
|  |  | Public | Private | Mixed | Public | Private | Mixed | Public | Private | Mixed |
| **\*Specimen collection capacity** | No capacity | 15(88.2%) | 6(85.7%) | 2(66.7%) | 12(52.2%) | 2(66.7%) | 1(50.0%) | 6(24.0%) | 9(42.9%) | 1(11.1%) |
|  | Inadequate | 0 | 1(14.3%) | 1(33.3%) | 3(13.0%) | 1(33.3%) | 1(50.0%) | 10(40.0%) | 9(42.9%) | 5(55.6%) |
|  | Partially adequate | 2(11.8%) | 0 | 0 | 4(17.4%) | 0 | 0 | 4(16.0%) | 2(9.5%) | 2(22.2%) |
|  | Adequate | 0 | 0 | 0 | 4(17.4%) | 0 | 0 | 5(20.0%) | 1(4.8%) | 1(11.1%) |
| **NP/throat swab kits** | Yes | 2(11.8%) | 0 | 0 | 8(34.8%) | 0 | 1(50.0%) | 13(52.0%) | 8(38.1%) | 3(33.3%) |
|  | No | 15(88.2%) | 7(100%) | 3(100%) | 15(65.2%) | 3(100%) | 1(50.0%) | 12(48.0%) | 13(61.9%) | 6(66.7%) |
| **VTM** | Yes | 0 | 0 | 1(33.3%) | 9(39.1%) | 0 | 1(50.0%) | 8(32.0%) | 1(4.8%) | 2(22.2%) |
|  | No | 17(100%) | 7(100%) | 2(66.7%) | 14(60.9%) | 3(100%) | 1(50.0%) | 17(68.0%) | 20(95.2%) | 7(77.8%) |
| **Portable fridge box** | Yes | 2(11.8%) | 0 | 0 | 7(30.4%) | 0 | 1(50.0%) | 5(20.0%) | 3(14.3%) | 2(22.2%) |
|  | No | 15(88.2%) | 7(100%) | 3(100%) | 16(69.6%) | 3(100%) | 1(50.0%) | 20(80.0%) | 18(85.7%) | 7(77.8%) |
| **Refrigerator** | Yes | 1(5.9%) | 2(28.6%) | 0 | 6(26.1%) | 0 | 0 | 6(24.0%) | 5(23.8%) | 2(22.2%) |
|  | No | 16(94.1%) | 5(71.4%) | 3(100%) | 17(73.9%) | 3(100%) | 2(100%) | 19(76.0%) | 16(76.2%) | 7(77.8%) |
| **Trained laboratory staff** | Yes | 1(5.9%) | 0 | 0 | 9(39.1%) | 0 | 1(50.0%) | 8(32.0%) | 5(23.8%) | 2(22.2%) |
|  | No | 16(94.1%) | 7(100%) | 3(100%) | 14(60.9%) | 3(100%) | 1(50.0%) | 17(68.0%) | 16(76.2%) | 7(77.8%) |
| **Transport mechanism** | Yes | 2(11.8%) | 0 | 0 | 5(21.7%) | 0 | 1(50.0%) | 7(28.0%) | 3(14.3%) | 0 |
|  | No | 15(88.2%) | 7(100%) | 3(100%) | 18(78.3%) | 3(100%) | 1(50.0%) | 18(72.0%) | 18(85.7%) | 9(100%) |
| **PCR** | Yes | 0 | 0 | 0 | 0 | 0 | 0 | 0 | 0 | 1(11.1%) |
|  | No | 17(100%) | 6(85.7%) | 3(100%) | 22(95.7%) | 3(100%) | 2(100%) | 23(92.0%) | 19(90.5%) | 7(77.8%) |
|  | Don't know | 0 | 1(14.3%) | 0 | 1(4.3%) | 0 | 0 | 2(8.0%) | 2(9.5%) | 1(11.1%) |
| **None** | Yes | 12(70.6%) | 5(71.4%) | 2(66.7%) | 9(39.1%) | 3(100%) | 1(50.0%) | 4(16.0%) | 8(38.1%) | 3(33.3%) |
|  | No | 5(29.4%) | 2(28.6%) | 1(33.3%) | 14(60.9%) | 0 | 1(50.0%) | 21(84.0%) | 13(61.9%) | 6(66.7%) |

\*No capacity (no service at all); Inadequate ( $\leq 50\%$ of those requiring service received it); Partially adequate ( $>50\%$ of those requiring service received it, but not all); Adequate (all of those requiring service received it)

Nasopharyngeal/throat swab kits were available in one-third (32%; 35/110) of hospitals. Large hospitals were found better equipped in this regard. However, viral transport media (VTM) were available in 20% (22/110) of hospitals only. Even the larger hospitals did not have adequate supply of VTM. Likewise, most of the hospitals (82%; 90/110) neither had portable fridge boxes, nor had the mechanism for specimen transport. Most (80%) of them lacked a refrigerator for storing specimens collected from COVID-19 suspects. Although 26 hospitals had a trained laboratory personnel who could process respiratory specimens for viral testing, only one hospital (large/tertiary) in the whole nation had a functional polymerase chain reaction (PCR) machine during the study period (Supplementary table 1).

### 3.5. Infection prevention and control measures

Simple face masks were in adequate supply in the majority of small (78%; 21/27), medium (86%; 24/28), and large (50/55) hospitals across the country. However, the number of hospitals with the supply of N95 respirator masks (5 small; 9 medium; 8 large) and Filter masks (2 each in small, medium, large) was low. Gloves were in adequate supply in the majority (88/110), but, there was a scarcity of eye and foot wears (Table 1).

Majority of small (24/27), medium (22/28), and large (49/55) hospitals did not have an arrangement of whole-body personal protective gears (PPEs) for health workers at the time of study. Public facilities had a relatively better availability of PPEs than non-public facilities: small:12% vs. 10%; medium:26% vs. none; large:16% vs. 7% (Table 1).

**Table 1.** Availability of infection prevention measures for health workers in small, medium, and large hospitals

|  |  | Type of Hospital (N=110) |  |  |  |  |  |  |  |  |
| --- | --- | --- | --- | --- | --- | --- | --- | --- | --- | --- |
|  |  | Small (n=27) |  |  | Medium (n=28) |  |  | Large (n=55) |  |  |
|  |  | Public | Private | Mixed | Public | Private | Mixed | Public | Private | Mixed |
| **Simple face mask** | Yes | 11(64.7%) | 7(100%) | 3(100%) | 19(82.6%) | 3(100%) | 2(100%) | 22(88.0%) | 20(95.2%) | 8(88.9%) |
|  | No | 6(35.3%) | 0 | 0 | 4(17.4%) | 0 | 0 | 3(12.0%) | 1(4.8%) | 1(11.1%) |
| **N95 mask** | Yes | 3(17.6%) | 1(14.3%) | 1(33.3%) | 8(34.8%) | 1(33.3%) | 0 | 3(12.0%) | 3(14.3%) | 2(22.2%) |
|  | No | 14(82.4%) | 6(85.7%) | 2(66.7%) | 15(65.2%) | 2(66.7%) | 2(100%) | 22(88.0%) | 18(85.7%) | 7(77.8%) |
| **Filter mask** | Yes | 0 | 2(28.6%) | 0 | 2(8.7%) | 0 | 0 | 1(4.0%) | 1(4.8%) | 0 |
|  | No | 17(100%) | 5(71.4%) | 3(100%) | 21(91.3%) | 3(100%) | 2(100%) | 24(96.0%) | 20(95.2%) | 9(100%) |
| **Gloves** | Yes | 14(82.4%) | 6(85.7%) | 3(100%) | 21(91.3%) | 2(66.7%) | 2(100%) | 16(64.0%) | 16(76.2%) | 8(88.9%) |
|  | No | 3(17.6%) | 1(14.3%) | 0 | 2(8.7%) | 1(33.3%) | 0 | 9(36.0%) | 5(23.8%) | 1(11.1%) |
| **Eye and foot wear** | Yes | 4(23.5%) | 0 | 1(33.3%) | 8(34.8%) | 1(33.3%) | 0 | 5(20.0%) | 5(23.8%) | 1(11.1%) |
|  | No | 13(76.5%) | 7(100%) | 2(66.7%) | 15(65.2%) | 2(66.7%) | 2(100%) | 20(80.0%) | 16(76.2%) | 8(88.9%) |
| **PPE whole body suit** | Yes | 2(11.8%) | 0 | 1(33.3%) | 6(26.1%) | 0 | 0 | 4(16.0%) | 2(9.5%) | 0 |
|  | No | 15(88.2%) | 7(100%) | 2(66.7%) | 17(73.9%) | 3(100%) | 2(100%) | 21(84.0%) | 19(90.5%) | 9(100%) |

Majority of the small, medium, and large hospitals had arrangements of hand sanitizers (21/27, 24/28, 43/55) and hand-washing bins (14/27, 22/28, 38/55) at each of the hospital units. Thermal gun was present in a half of the total hospitals under study (small:7/27; medium:18/28; large:29/55; overall 54/110). The availability of thermal gun was relatively higher in public sector than in non-public for small (35% vs. 10%) and medium hospitals (65% vs. 60%), and in non-public sector than in public for larger hospitals (60% vs. 44%) (Table 2).

Nearly half of the facilities had set up ‘Health Information Desk’ within their premises as a response to COVID-19 pandemic: 33% (9/27) of small, 50% (14/28) of medium, and 40% (22/55) of large hospitals. One-fourth (24%; 27/110) had adopted disinfection techniques, whereas one-fifth (19%; 21/110) had a proper waste disposal mechanism (Table 2).

**Table 2.** Availability of infection prevention and control measures in hospitals

|  |  | Type of Hospital (N=110) |  |  |  |  |  |  |  |  |
| --- | --- | --- | --- | --- | --- | --- | --- | --- | --- | --- |
|  |  | Small (n=27) |  |  | Medium (n=28) |  |  | Large (n=55) |  |  |
|  |  | Public | Private | Mixed | Public | Private | Mixed | Public | Private | Mixed |
| **Hand sanitizers** | Yes | 13(76.5%) | 5(71.4%) | 3(100%) | 20(87.0%) | 2(66.7%) | 2(100%) | 20(80.0%) | 15(71.4%) | 8(88.9%) |
|  | No | 4(23.5%) | 2(28.6%) | 0 | 3(13.0%) | 1(33.3%) | 0 | 5(20.0%) | 6(28.6%) | 1(11.1%) |
| **Hand washing basins** | Yes | 9(52.9%) | 2(28.6%) | 3(100%) | 18(78.3%) | 2(66.7%) | 2(100%) | 17(68.0%) | 15(71.4%) | 6(66.7%) |
|  | No | 8(47.1%) | 5(71.4%) | 0 | 5(21.7%) | 1(33.3%) | 0 | 8(32.0%) | 6(28.6%) | 3(33.3%) |
| **Thermal Gun** | Yes | 6(35.3%) | 0 | 1(33.3%) | 15(65.2%) | 2(66.7%) | 1(50.0%) | 11(44.0%) | 14(66.7%) | 4(44.4%) |
|  | No | 11(64.7%) | 7(100%) | 2(66.7%) | 8(34.8%) | 1(33.3%) | 1(50.0%) | 14(56.0%) | 7(33.3%) | 5(55.6%) |
| **Health Information desk** | Yes | 7(41.2%) | 0 | 2(66.7%) | 12(52.2%) | 1(33.3%) | 1(50.0%) | 10(40.0%) | 6(28.6%) | 6(66.7%) |
|  | No | 10(58.8%) | 7(100%) | 1(33.3%) | 11(47.8%) | 2(66.7%) | 1(50.0%) | 15(60.0%) | 15(71.4%) | 3(33.3%) |
| **Disinfection technique** | Yes | 4(23.5%) | 2(28.6%) | 0 | 3(13.0%) | 0 | 1(50.0%) | 7(28.0%) | 7(33.3%) | 3(33.3%) |
|  | No | 13(76.5%) | 5(71.4%) | 3(100%) | 20(87.0%) | 3(100%) | 1(50.0%) | 18(72.0%) | 14(66.7%) | 6(66.7%) |
| **Waste disposal system** | Yes | 3(17.6%) | 1(14.3%) | 1(33.3%) | 5(21.7%) | 0 | 2(100%) | 4(16.0%) | 3(14.3%) | 2(22.2%) |
|  | No | 14(82.4%) | 6(85.7%) | 2(66.7%) | 18(78.3%) | 3(100%) | 0 | 21(84.0%) | 18(85.7%) | 7(77.8%) |

### 3.6. Human resource for pandemic response

None of the small hospitals had an Infectious disease (ID) physician, a microbiologist, or a nurse trained on infectious disease (Table 3). Only one out of 28 medium hospitals had a range of staff providing health service: ID physician (private), microbiologist (public), public health specialist (mixed), and researcher (mixed); five medium hospitals (3 public, 2 private) had a trained nurse. Majority of small and medium hospitals did not have a molecular biologist (laboratory technologist) for collecting and processing patient specimens.

Except for General practitioners (MDGPs)—available in over half of the facilities (30/55), all other human resource for health were lacking mostly in large hospitals (Table 3).

**Table 3.** Human resource for health available in hospitals

|  |  | Type of Hospital (N=110) |  |  |  |  |  |  |  |  |
| --- | --- | --- | --- | --- | --- | --- | --- | --- | --- | --- |
|  |  | Small (n=27) |  |  | Medium (n=28) |  |  | Large (n=55) |  |  |
|  |  | Public | Private | Mixed | Public | Private | Mixed | Public | Private | Mixed |
| **Infectious disease physician** | Yes | 0 | 0 | 0 | 0 | 1(33.3%) | 0 | 5(20.0%) | 3(14.3%) | 2(22.2%) |
|  | No | 17(100%) | 7(100%) | 3(100%) | 23(100%) | 2(66.7%) | 2(100%) | 20(80.0%) | 18(85.7%) | 7(77.8%) |
| **MDGP** | Yes | 0 | 2(28.6%) | 2(66.7%) | 16(69.6%) | 3(100%) | 2(100%) | 12(48.0%) | 13(61.9%) | 5(55.6%) |
|  | No | 17(100%) | 5(71.4%) | 1(33.3%) | 7(30.4%) | 0 | 0 | 13(52.0%) | 8(38.1%) | 4(44.4%) |
| **Nurse trained on Infectious disease** | Yes | 0 | 0 | 0 | 3(13.0%) | 2(66.7%) | 0 | 3(12.0%) | 4(19.0%) | 3(33.3%) |
|  | No | 17(100%) | 7(100%) | 3(100%) | 20(87.0%) | 1(33.3%) | 2(100%) | 22(88.0%) | 17(81.0%) | 6(66.7%) |
| **Molecular laboratory technologist** | Yes | 1(5.9%) | 0 | 0 | 1(4.3%) | 0 | 1(50.0%) | 5(20.0%) | 7(33.3%) | 4(44.4%) |
|  | No | 16(94.1%) | 7(100%) | 3(100%) | 22(95.7%) | 3(100%) | 1(50.0%) | 20(80.0%) | 14(66.7%) | 5(55.6%) |
| **Microbiologist** | Yes | 0 | 0 | 0 | 1(4.3%) | 0 | 0 | 0 | 2(9.5%) | 1(11.1%) |
|  | No | 17(100%) | 7(100%) | 3(100%) | 22(95.7%) | 3(100%) | 2(100%) | 25(100%) | 19(90.5%) | 8(88.9%) |
| **Public health specialist** | Yes | 0 | 0 | 0 | 0 | 0 | 1(50.0%) | 5(20.0%) | 1(4.8%) | 3(33.3%) |
|  | No | 17(100%) | 7(100%) | 3(100%) | 23(100%) | 3(100%) | 1(50.0%) | 20(80.0%) | 20(95.2%) | 6(66.7%) |
| **Researcher** | Yes | 0 | 0 | 0 | 0 | 0 | 1(50.0%) | 4(16.0%) | 3(14.3%) | 4(44.4%) |
|  | No | 17(100%) | 7(100%) | 3(100%) | 23(100%) | 3(100%) | 1(50.0%) | 21(84.0%) | 18(85.7%) | 5(55.6%) |
| **None** | Yes | 16(94.1%) | 4(57.1%) | 1(33.3%) | 6(26.1%) | 0 | 0 | 5(20.0%) | 5(23.8%) | 2(22.2%) |
|  | No | 1(5.9%) | 3(42.9%) | 2(66.7%) | 17(73.9%) | 3(100%) | 2(100%) | 20(80.0%) | 16(76.2%) | 7(77.8%) |

### 3.7. Stock of (essential) antiviral drugs

None of the small and medium (except one) hospitals had a stock of Oseltamivir which is an antiviral drug considered effective against influenza-like respiratory illness. Only five (9%) of the large hospitals had this drug available for dispensing. Approximately half of the hospitals had a stock of Chloroquine/Hydroxychloroquine(HC) which was rampantly used (for both therapeutic and research purposes) globally during study period.

Remdesivir, an antiviral drug under clinical trials (recently approved by the Food and Drug Authority or FDA, USA for COVID-19 treatment) was available in only one hospital (large, public) out of 110 hospitals studied (Supplementary table 2).

**Supplementary Table 2.** Stock of potentially useful antiviral medicines in the hospitals

|  |  | Type of Hospital |  |  |  |  |  |  |  |  |
| --- | --- | --- | --- | --- | --- | --- | --- | --- | --- | --- |
|  |  | Small |  |  | Medium |  |  | Large |  |  |
|  |  | Public | Private | Mixed | Public | Private | Mixed | Public | Private | Mixed |
| **Oseltamivir** | Yes | 0 | 0 | 0 | 1(4.3%) | 0 | 0 | 3(12.0%) | 2(9.5%) | 0 |
|  | No | 17(100%) | 7(100%) | 3(100%) | 22(95.7%) | 3(100%) | 2(100%) | 22(88.0%) | 19(90.5%) | 9(100%) |
| **Remdesivir** | Yes | 0 | 0 | 0 | 0 | 0 | 0 | 1(4.0%) | 0 | 0 |
|  | No | 17(100%) | 7(100%) | 3(100%) | 23(100%) | 3(100%) | 2(100%) | 24(96.0%) | 21(100%) | 9(100%) |
| **Chloroquine/HC** | Yes | 6(35.3%) | 3(42.9%) | 1(33.3%) | 7(30.4%) | 1(33.3%) | 1(50.0%) | 16(64.0%) | 9(42.9%) | 5(55.6%) |
|  | No | 11(64.7%) | 4(57.1%) | 2(66.7%) | 16(69.6%) | 2(66.7%) | 1(50.0%) | 9(36.0%) | 12(57.1%) | 4(44.4%) |
| **None** | Yes | 11(64.7%) | 4(57.1%) | 2(66.7%) | 15(65.2%) | 2(66.7%) | 1(50.0%) | 9(36.0%) | 12(57.1%) | 4(44.4%) |
|  | No | 6(35.3%) | 3(42.9%) | 1(33.3%) | 8(34.8%) | 1(33.3%) | 1(50.0%) | 16(64.0%) | 9(42.9%) | 5(55.6%) |

### 3.8. Government’ support and intersectoral engagement

Majority of the hospitals had not received COVID-19 related technical or financial support from the government until the second week of lockdown (7^th^ April), although such support was expected as and when the WHO declared the disease as a global pandemic on 11^th^ March, 2020 (Table 4).

Optimization of case reporting and referral mechanism was also poorly performed across the facilities—only two small, five medium, and ten large hospitals supported for case referral; three small and 13 medium, and 15 large hospitals for case reporting. Public facilities were comparatively better supported than private facilities in terms COVID-19 HEIC materials, (35%; 23/65 vs. 26%; 8/31) (Table 4).

**Table 4.**
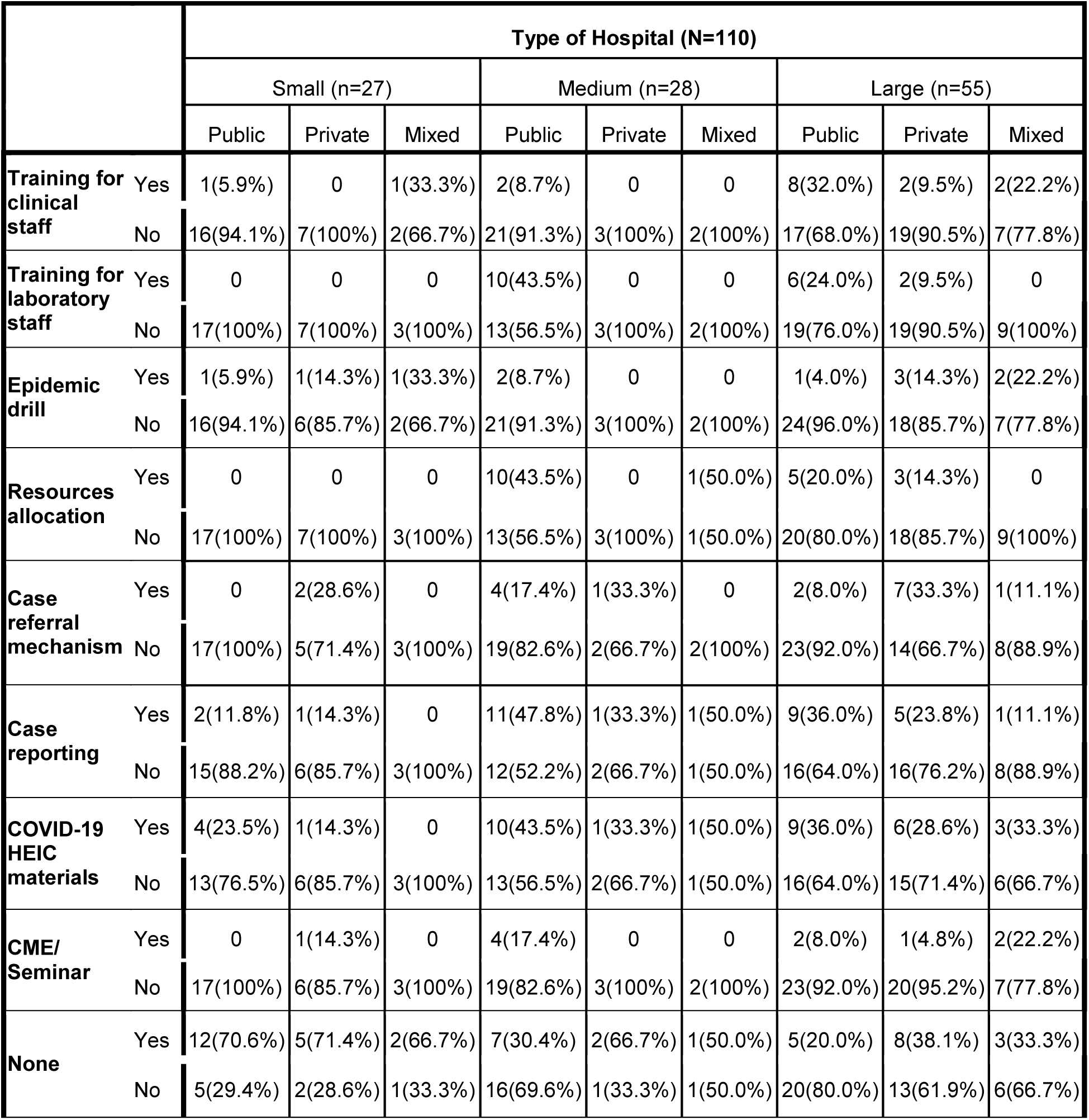
Distribution of Government’s pandemic support to hospitals, by hospital type

### 3.9. Province-wise service availability

By 7^th^ April, more than half of the health facilities in each province had allocated at least one isolation bed for COVID-19 suspected or confirmed patient. However, only few hospitals had arranged up to 20 isolation beds: one in provinces 1, Karnali, and Sudurpaschim; two in Province 5; four in Gandaki; and six in Bagmati. Three hospitals in Province 1, two in Bagmati, and one each in Gandaki and Province 5 had arranged >20 isolation beds (Figure 5A).

Majority of the hospitals in each province did not have a functional ICU bed for critical care service. Only a few hospitals in Province 1 (2/18), Bagmati (3/33), and Gandaki (2/21) had better ICU service with >16 beds (Figure 5B).

**Figure 5.**
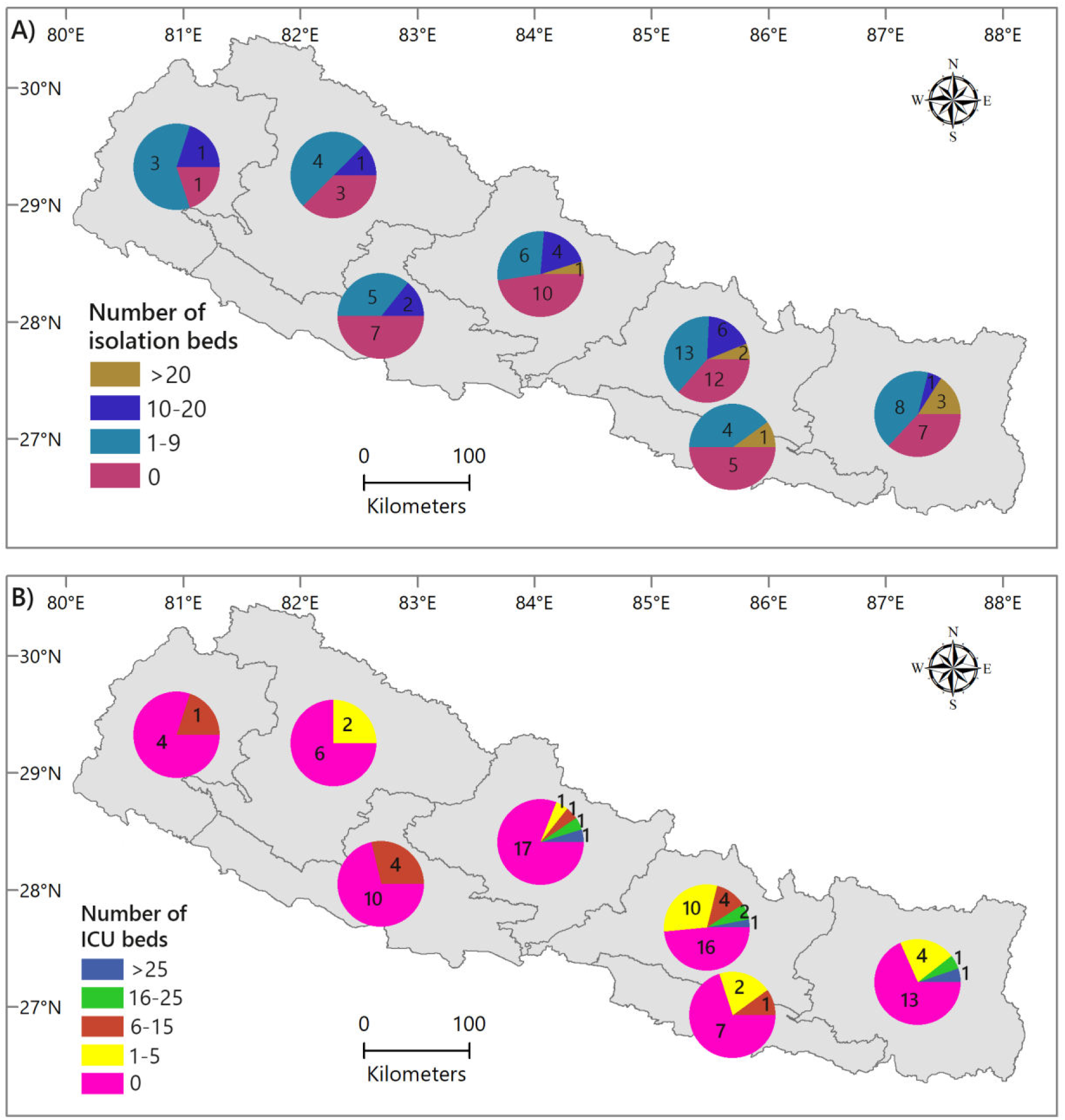
Availability of isolation beds (A) and functional ICU beds (B) for COVID-19 suspected patients in hospitals, by province.

Thermal gun for rapid temperature check was available in 68% of facilities in Province 1; 54% in Bagmati; and 60% in Sudurpaschim. Viral transport media (VTM), an important equipment for lodging and transporting respiratory specimens, was lacking in the majority of facilities across the provinces, with Province 2 and 5 reporting availability in only one hospital. Similar scarcity was reported in terms of full protective gears (PPEs) for health workers. Majority (75-91%) of the hospitals across seven provinces reported unavailability of PPE suits (Figure 6).

**Figure 6.**
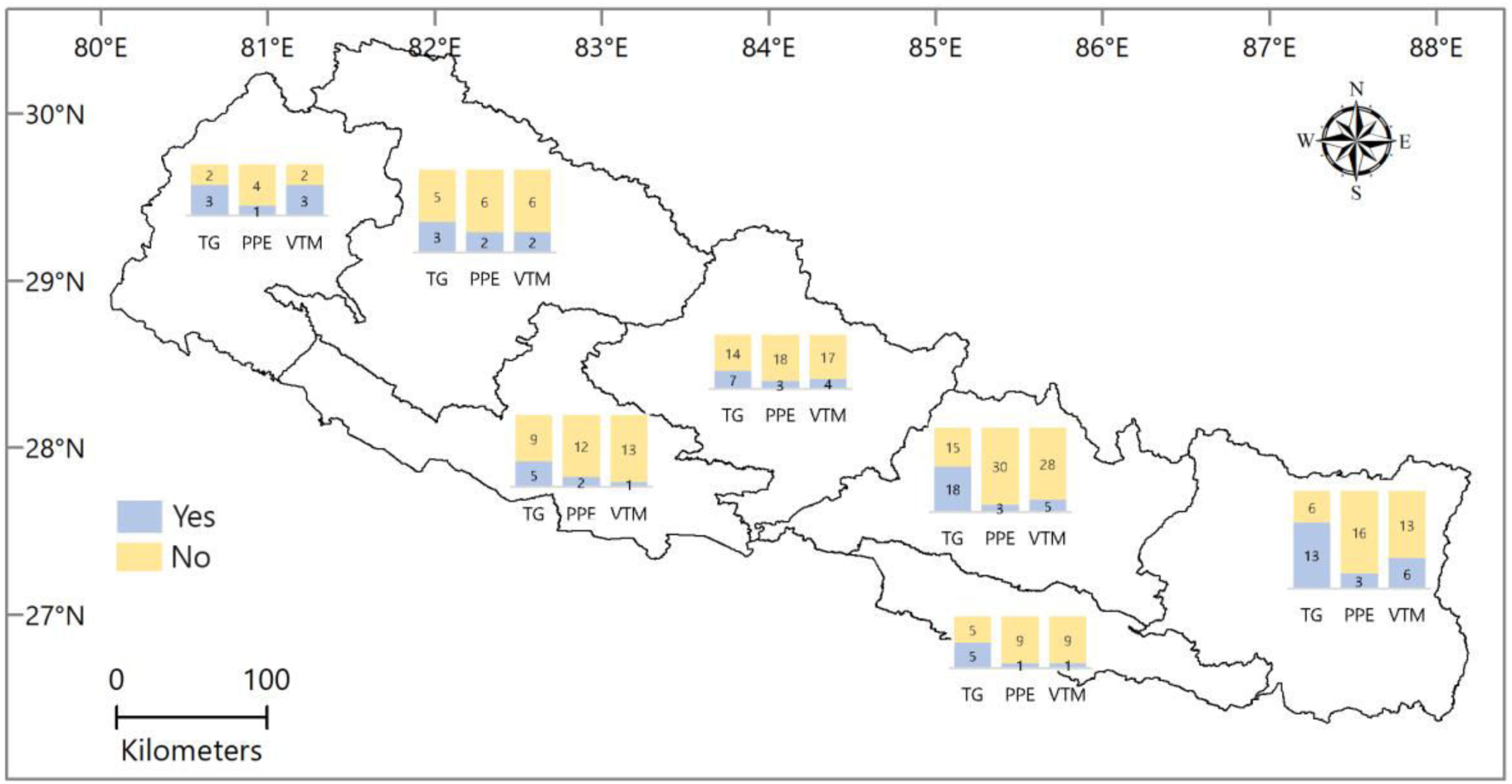
Availability of thermal gun, personal protective equipment, and viral transport media in hospitals, by province.

Approximately one-third of health facilities in Province 1 and Karnali received government’s support for clinical and laboratory training, whereas less or no facilities in other provinces reported, clinical such opportunity. Allocation of resources—equipment, emergency fund, personal protective gears, was relatively better in Province 1, Bagmati, Karnali and Sudurpaschim; on an average, one-fourth of the facilities reported the receipt. However, allocation of health education and information (HEIC) resources was evenly poor in all provinces. Province 2 did not receive government’s support for the optimization of COVID-19 case reporting and referral mechanism, whereas such support reached up to 20% of health facilities in Province 5, and relatively more facilities (up to one-third) in the remaining provinces (Supplementary table 3).

**Supplementary Table 3.** Government’s pandemic support to hospitals, by provinces

|  |  | Province |  |  |  |  |  |  |
| --- | --- | --- | --- | --- | --- | --- | --- | --- |
|  |  | Province 1 | Province 2 | Bagmati | Gandaki | Province 5 | Karnali | Sudurpaschim |
| **Training for clinical staff** | Yes | 1(5.3%) | 0 | 9(27.3%) | 4(19.0%) | 2(14.3%) | 0 | 0 |
|  | No | 18(94.7%) | 10(100%) | 24(72.7%) | 17(81.0%) | 12(85.7%) | 8(100%) | 5(100%) |
| **Training for lab staff** | Yes | 6(31.6%) | 1(10.0%) | 4(12.1%) | 4(19.0%) | 0 | 3(37.5%) | 0 |
|  | No | 13(68.4%) | 9(90.0%) | 29(87.9%) | 17(81.0%) | 14(100%) | 5(62.5%) | 5(100%) |
| **Epidemic drill** | Yes | 0 | 0 | 6(18.2%) | 5(23.8%) | 0 | 0 | 0 |
|  | No | 19(100%) | 10(100%) | 27(81.8%) | 16(76.2%) | 14(100%) | 8(100%) | 5(100%) |
| **Resources allocation** | Yes | 4(21.1%) | 1(10.0%) | 8(24.2%) | 2(9.5%) | 0 | 2(25.0%) | 2(40.0%) |
|  | No | 15(78.9%) | 9(90.0%) | 25(75.8%) | 19(90.5%) | 14(100%) | 6(75.0%) | 3(60.0%) |
| **Referral mechanism** | Yes | 4(21.1%) | 0 | 4(12.1%) | 4(19.0%) | 2(14.3%) | 2(25.0%) | 1(20.0%) |
|  | No | 15(78.9%) | 10(100%) | 29(87.9%) | 17(81.0%) | 12(85.7%) | 6(75.0%) | 4(80.0%) |
| **Case reporting** | Yes | 7(36.8%) | 0 | 10(30.3%) | 7(33.3%) | 1(7.1%) | 2(25.0%) | 4(80.0%) |
|  | No | 12(63.2%) | 10(100%) | 23(69.7%) | 14(66.7%) | 13(92.9%) | 6(75.0%) | 1(20.0%) |
| **Information/HIEC materials** | Yes | 7(36.8%) | 2(20.0%) | 7(21.2%) | 6(28.6%) | 5(35.7%) | 5(62.5%) | 3(60.0%) |
|  | No | 12(63.2%) | 8(80.0%) | 26(78.8%) | 15(71.4%) | 9(64.3%) | 3(37.5%) | 2(40.0%) |
| **COVID-related CME/Seminar** | Yes | 1(5.3%) | 1(10.0%) | 2(6.1%) | 3(14.3%) | 1(7.1%) | 1(12.5%) | 1(20.0%) |
|  | No | 18(94.7%) | 9(90.0%) | 31(93.9%) | 18(85.7%) | 13(92.9%) | 7(87.5%) | 4(80.0%) |
| **None** | Yes | 8(42.1%) | 6(60.0%) | 15(45.5%) | 7(33.3%) | 5(35.7%) | 3(37.5%) | 1(20.0%) |
|  | No | 11(57.9%) | 4(40.0%) | 18(54.5%) | 14(66.7%) | 9(64.3%) | 5(62.5%) | 4(80.0%) |

## 4. DISCUSSION

The overall service availability including specimen collection and laboratory services, isolation of COVID-19 suspects or cases, and ICU bed was found to be severely inadequate across small, medium, and large hospitals at the time of the study. Although simple IPC measures such as gloves, simple face masks, hand sanitizers and hand-washing basins were adequately available, there was a lack of proper disinfection and waste disposal mechanism. Medium hospitals (district/community hospitals) had better supply of N95 masks and PPE suits than small (PHCs) and large hospitals. Additionally, human resource for pandemic was found inadequate across all provinces, so was the government’s COVID-19 support in terms of training, case reporting and referral, resources, and risk communication strategy. Similar service constraints were reported by the National Disaster Risk Reduction Center.(13) The United Nations Development Programme (UNDP) COVID-19 and Human Development Report 2020 also confirmed low preparedness to COVID-19 in Nepal, measured on the basis of national human development index (HDI), HDI inequalities, and health systems capacity.(14)

By the time the survey was closed (7^th^ April 2020), i.e. a month after the WHO declared COVID-19 as global pandemic, a massive number of health facilities in many high-income countries had been affected by daily case surge with high rates of mortality. In contrast, Nepal had detected only nine cases with less than 1,000 PCR tests performed, and the government was stumbling through the second week of nationwide lockdown. Despite that given window of opportunity for Nepal in terms of pandemic preparedness, the majority of small and over one-third of medium and large hospitals had not allocated isolation beds for COVID-19 suspects. Even a number of large (tertiary care) facilities (50%) did not have a functional ICU unit; and among those with such availability, 71% had five or less functional beds that could be offered to COVID-19 patients if needed. With the decision HLCC decision (17^th^ March) to expand isolation and ICU beds throughout the country, a ray of hope spread amongst the public. (9) However, it is still unknown how the decision was implemented. Performance of the concerned ministries and health facilities while implementing decisions was often poor in Nepal. However, the concerned ministries and health facilities performed poorly in the implementation aspect. It should be noted that many countries utilized large halls, unused buildings, parks or open spaces to set up new isolation centers, but the scarcity of ICU beds was haunting everyone regardless of the country’s economy.

Prior to and in the first two weeks of lockdown, the majority of the hospitals were seeing a number of patients with respiratory (COVID-like) symptoms; some large hospitals reporting >200 COVID-19 suspected patients per day. As the hospitals started collecting respiratory specimens, only a few of them had an adequate laboratory capacity. Four out of five hospitals did not have a supply of VTM and just one large hospital had a functional PCR machine throughout the country. The majority of small and medium hospitals did not have a molecular biologist (laboratory technologist) for collecting and processing specimens. Although the Government of Nepal stipulates that each health facility should have its own laboratory and a pharmacy, many hospitals had to rely on the private laboratories doing business in the periphery. In the present context, PCR-based testing has been expanded to all seven provinces, still, there are fresh reports of the scarcity of VTM and test reagents.

Face masks and gloves were in adequate supply in the majority of hospitals, but there was a gross lack of N95 respirators, Filter masks, eye and foot wears, and whole-body PPE suits for healthcare workers. Despite IPC guidance issued by Nepal Medical Council and the Health Emergency Operations Center (HEOC) for providing care to COVID-19 suspected cases, clinicians throughout the country expressed constant fears and concerns of potential exposures in various social media platforms.(15)(16) Thermal guns that measure patient’s temperature were better available in non-public hospitals than in public; one reason could be a relatively quicker procurement process in non-public hospitals as compared to sluggish procurement in government facilities even during the emergencies. The highest number of facilities with Thermal gun was in Province 1 (68%) and Sudurpaschim (60%), the lowest in Gandaki (33%). This difference could be explained by the close proximity of former two provinces to India with an easy cross-border import.

According to the Center for Disease Control and Prevention (CDC, USA), the planning, preparedness and response to a pandemic is a team work.(17) It involves health care workers, public health professionals, researchers, scientists, politicians, private sector, community, and individual experts working together to solve a common problem. In this study, human resource for COVID-19 response was found inadequate across the range of facilities. Although a qualified General practitioner (MDGP) was providing service in more than half of medium and large hospitals, other health cadres crucial during pandemic response— physicians and nurses trained in Infectious disease, microbiologists, public health specialists, and clinical epidemiologists, were on board in less than 20% of hospitals. According to the UNDP report, Nepal’s health system capacity—measured on the basis of human resource for health, is very low with the availability of just three hospital beds, six physicians, and 27 nurses per 100,000 people.(14) As Nepal prepares to tackle community outbreaks now and in the future, it is important for both public and private sectors to invest more on human resource should they expect better health outcomes and improved national health indicators.

Province wise, the government’s support to the health facilities was not satisfactory at the time of this study. Allocation of needful resources such as technical personnel, laboratory equipment, personal protective gears for health workers, health education (HEIC) materials, and emergency fund, which were the minimum expectations from the Department of Health Services (DoHS) at the time of health emergencies, was absent in all small hospitals, patchy in medium (district hospitals), and negligible in large hospitals. Provinces 2 and 5 were less supported than other provinces in terms of resource allocation, technical support, and optimization of COVID-19 case report and referral mechanisms. This deficit has been reflected by more rapid rise in cases in Province 2 (96 cases) and 5 (70 cases) than other provinces.(6) Much needed epidemic drill was also lacking throughout provinces, except Bagmati (six drills conducted) and Gandaki (five drills conducted). Other special services such as isolation beds for COVID-19 suspects, functional ICU beds for critically ill patients, sample collection and testing capacity, as well as, the level of infection prevention and control, was overall poor in all provinces. Our findings are supported by the field studies conducted by the National Disaster Risk Reduction Center.(13)(18)

To mitigate pandemic-borne workload and managerial confusion within the Health Service divisions, the government could have leveraged full authority to the provinces and municipals, coupled with stringent action plans for high priority tasks—procurement of equipment and supplies, recruitment and training of human resource, expansion of laboratories and hospital units, contact tracing, and case reporting. On the other hand, the healthcare facilities, small to large and public or private, could have proactively activated their ‘emergency preparedness plan’ to ease the operational processes, way before the disease dismantled local health systems. Utilizing the relatively larger window of preparedness opportunity compared to hard-hit countries, these facilities could have adopted full preparation gears early on adhering to the relevant national guidelines and service standards, for example, Hospital Management Strengthening Program (HMSP)—Minimum Service Standards (MSS) checklist for hospitals, Nepal Health Infrastructure Development Standards (HIDS).(19)(20)

With the above findings, it is worthwhile to mention few limitations of this study. First, it was conducted using internet-based tool considering the government’s strict orders for travel restriction and infection prevention. A field-based observational study would better reflect the scenario of services and IPC measures adopted by the hospitals. Second, the study did not cover all hospitals of the country; only a few-select hospitals could be included. Third, the study took the perspectives of frontline physicians only, excluding other cadres of health workforce (nurses, paramedics) who were also involved in the response.

## 5. Conclusions and recommendations

Despite the above-mentioned limitations, our study found inadequacy in several aspects of health services and IPC measures that define hospital readiness in the context of COVID-19 case surge. We also found that the government’s pandemic response or support was unevenly distributed across the provinces and health facilities of Nepal. As this pandemic teaches an important lesson for all governments—to act early and proactively during health emergencies and not wait until the disease disrupts health systems, our study findings could provide them further guidance to formulate emergency preparedness strategic plans and reduce the impact. Furthermore, to know the actual pandemic preparedness at each service level, and to understand the exact scenarios of case surge and patient management, surveillance and reporting, essential health services, and logistic supplies, there is an urgent need to conduct large-scale studies, coordinated either by government’s epidemiology and research divisions or by non-government development and implementing partners. From global health point of view, other countries at similar levels of economic development could learn from our findings and conduct similar assessments in order to gauge their pandemic response.

## Data Availability

All data related to this research has been included in the manuscript and supplementary files. Additional details will be available upon special request to the main author.

## Acknowledgement

Researchers would like to acknowledge clinicians and health facilities that participated in this survey, and independent anonymous reviewers who provided critical feedback. Uttam Babu Shrestha is supported by National Geographic Society Grant (NGS-62058R-19).

## Conflict of interest

**None**

## Funding

**None**

## Notes

### Competing Interest Statement

The authors have declared no competing interest.

### Funding Statement

No funding received for this work.

